# Association between sociodemographic factors, visual acuity and corneal topographic outcomes after collagen cross-linking in patients with keratoconus

**DOI:** 10.1101/2025.08.26.25334513

**Authors:** Cara Heppell, Paul Nderitu, Ruchi Gour, Sophie Jones

**Affiliations:** King’s College Hospital NHS Foundation Trust, London, SE5 9RS, UK; Section of Ophthalmology, Faculty of Life Sciences and Medicine, King’s College London, London, WC2R 2LS, UK

## Abstract

**Purpose:** To determine the association between sociodemographic factors, visual acuity (VA) and corneal topography (Kmax and K2) following collagen cross-linking (CXL) in a cohort of ethnically-diverse keratoconus patients.

**Methods:** The records of 88 keratoconus patients who underwent CXL between January 2021 and December 2022 at King’s College Hospital were examined. Data on age, sex, ethnicity, deprivation deciles, significant postoperative complications, pre and postoperative best VA (BVA), Kmax and K2 were extracted. Univariate Kaplan-Meier and multivariate cox-regression survival analyses were used to determine outcomes of BVA stability, Kmax stability K2 stability and composite Kmax/K2 stability at 52 weeks after CXL.

**Results:** At 52 weeks, there was an 81.1% (95% confidence interval (CI): 71.2%–87.6%) probability of BVA stability and 80.7% (67.4%–88.5%) probability of Kmax/K2 stability. Multivariate analyses showed significant associations (*p*<0.05) between Kmax/K2 stability and ‘other’ ethnicity but not other sociodemographic factors.

**Conclusion:** CXL is associated with the stabilisation of vision and corneal topography in the majority of eyes. One significant association was found between other ethnicity and corneal topographical outcomes after CXL, however, no other significant associations were found between the majority of sociodemographic or economic factors with VA or corneal topographical outcomes after CXL. This suggests CXL results in good functional and anatomical outcomes independent of sociodemographic status.

## INTRODUCTION

Keratoconus is characterised by bilateral, asymmetrical, progressive non-inflammatory thinning and steepening of the cornea resulting in irregular astigmatism and decreased visual acuity [1-3] with onset most often in the second and third decades of life [4, 5]. The global prevalence of keratoconus is estimated to be 1.38 per 1000, with an increased incidence amongst black, Latino and Asian ethnicities [6, 7]. A retrospective cohort study in the United States identified an odds ratio (OR) of 1.57 and 1.43 for black and Latino patients respectively of being diagnosed with keratoconus compared to white patients, whereas an OR of 0.61 was identified for Asian patients [7]. However, a retrospective cohort study in England showed a fourfold increase in Asian patients compared with white patients, alongside earlier presentation and faster deterioration [8].

The pathogenesis of keratoconus is unknown, but several risk factors include eye rubbing, family history, allergy, asthma, and eczema [6]. Quality of life (QoL) is reduced in these patients, causing functional, symptomatic and psychological effects, which continue to decline over time [9, 10]. A best corrected visual acuity (VA) better than 0.3 logarithm of the minimum angle of resolution (LogMAR) or treatment with corneal cross-linking (CXL) have been associated with higher QoL scores [11]. Depending on the stage and stability of the condition, the management of keratoconus includes contact lenses, corneal cross-linking (CXL), intracorneal ring segment implantation (ICRS) and keratoplasty [12].

CXL involves the use of riboflavin and ultraviolet-A (UVA) light to crosslink collagen covalently, thereby stabilising the cornea [13], in which stiffening and flattening have been identified postoperatively [13-16]. The efficacy of CXL has been demonstrated in randomised control trials (RCTs) [16-21], which show long-term delay of progression and improvement in corneal topography measurements and VA [19, 22], thus reducing the need for corneal transplantation [23, 24]. Due to the risk of complications, including a reported 2.9% risk of reduced VA and 7.6% risk of continued progression postoperatively [14], CXL is offered to patients with unstable keratoconus as evidenced by disease progression usually determined by corneal topographic measurements and refraction [25].

Studies have demonstrated an association between sociodemographic factors and increased keratoconus severity, corneal transplantation and loss to follow up [26-29]. However, to our knowledge, there are no studies which have investigated the association between sociodemographic factors, VA and changes in corneal topography after CXL. Given the impact of the disease on the QoL of young, working age adults, any respective associations could have important implications for clinical management, patient selection or patient education.

In this study within an ethnically-diverse cohort of keratoconus patients, we used univariate and multivariate survival analyses to determine the association between sociodemographic factors of age, sex, ethnicity and socioeconomic deprivation and important outcomes of CXL treatment, namely stability of VA and corneal topography (Kmax and K2). Socioeconomic deprivation was measured using the deciles for income deprivation, employment and education deprivation. The deprivation indices are subdomains of the index of multiple deprivation (IMD), which is an aggregate measure of relative neighbourhood-level deprivation, whereby small geographical areas are ranked nationally into deciles deprivation [30].

## METHODS

### Data collection and study population

In a single tertiary centre (Kings College Hospital, Denmark Hill), an audit was conducted on local outcomes in all keratoconus patients who underwent CXL treatment between 1^st^ January 2021 to 31^st^ December 2022 then followed-up to the 31^st^ March 2024 using the published literature as the gold standard [22]. The data was accessed and extracted between the 9th March 2024 until the 16th December 2024. All data was anonymised by frontline clinical staff at extraction and study researchers had access to anonymised data only. Anonymised data included age, sex, ethnicity and deciles of IMD, either combined or for the subdomains of income, employment and education. Additionally, eye laterality, pre- and post-operative VA, Kmax, K2 and significant postoperative complications (e.g. infective corneal infiltrates) were obtained from electronic medical records (Medisoft) and corneal topography (Pentacam). This study was conducted in compliance with the tenets of the Declaration of Helsinki.

### Outcomes

The main post-operative outcomes were (1) Best VA (BVA: best of pinhole, unaided or corrected VA) stability, defined as 0.0 logMAR or less change from baseline, (2) Kmax/K2 stability, defined by 0.00 diopter (D) or less change from baseline in either Kmax or K2. Kmax and K2 stability outcomes were also analysed individually with results shown in the **Supporting Information**. The BVA stability threshold was set at 0.00 logMAR and Kmax/K2 stability at 0.00D to assess the risk of patients having functional worsening in VA or structural worsening on corneal topography after CXL from their pre-operative baseline.

### Statistical analysis

Descriptive statistics were summarised using means and standard deviations (SD) or counts and percentages. Univariate Kaplan-Meier curves with associated risk tables were created for each of the outcomes (**S1-4 Figs**.). Cox-regression survival analyses were performed to investigate the association between the co-variates of age, sex, ethnicity, socioeconomic deprivation (income, employment and education), preoperative Kmax, significant postoperative complications with each of the post-operative outcomes. Multivariate cox-regression models were adjusted for preoperative Kmax only, as a measure of corneal topography at baseline, since this measure also strongly correlates with preoperative K2 (R^2^=0.80). Separate Cox-regression analyses were run for income, employment, and education deprivation subdomains as the variables are strongly correlated and cannot be modelled concurrently. Patients with missing data for postoperative outcomes were excluded from survival analyses (**Fig 1**). Only one eye (first operated eye) was included per patient. R [31], and R Studio [32] were used to analyse the data (see **Supporting Information** for code used for each R analysis).

**Fig 1.**
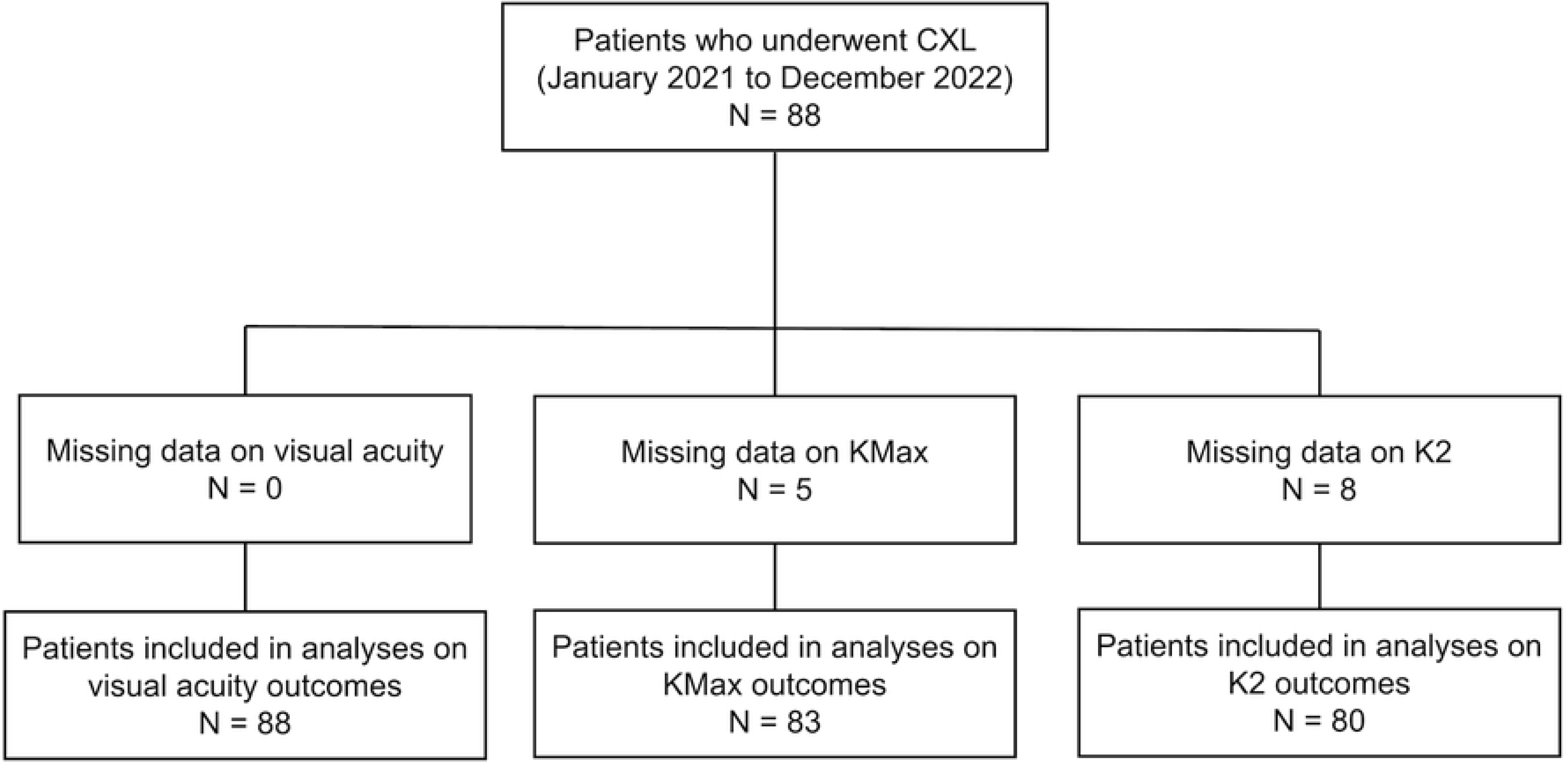
Flow chart of eyes included in data analyses. *CXL:* corneal cross linking.

## RESULTS

A total of 88 keratoconus patients underwent CXL between January 2021 and December 2022. The mean age was 24.6 years (SD: 6.8), 63.6% were male and 55.7% were left eyes (**Table 1**). The majority of patients were of black (40.9%) or white (23.9%) ethnicity. Mean aggregate IMD, income, employment, and education decile were 3.9 (SD 1.9), 3.8 (SD 2.1), 4.4 (SD 2.1) and 5.2 (SD 2.1) respectively. Deciles for aggregate IMD, and deprivation subdomains varied across ethnicity groups (**Fig 2**). Patients from a white ethnic background had the highest mean overall IMD (least deprived, 4.4) and employment decile (4.8). Patients of mixed ethnicity had highest mean income deprivation decile (4.3), whilst patients of black ethnicity had the highest mean education deprivation decile (6.1). Patients from the ‘other’ ethnicity group had the lowest mean overall (most deprived), IMD, income, employment and education deprivation deciles (3.0, 3.3, 3.7 and 2.7 respectively).

**TABLE 1.** COHORT CHARACTERISTICS

|  | **Mean (SD)**<br>**or N [%]** | **Missing** |
| --- | --- | --- |
| **Age** | 24.6 (6.8) | 0 |
| **Sex** |  |  |
| **Male** | 56 [63.6%] | - |
| **Female** | 32 [36.4%] | - |
| **Ethnicity** |  |  |
| **White** | 21 [23.9%] | - |
| **Black** | 36 [40.9%] | - |
| **South Asian** | 5 [5.7%] | - |
| **Mixed** | 3 [3.4%] | - |
| **Other** | 3 [3.4%] | - |
| **Not Specified** | 20 [22.7%] | - |
| **IMD** | 3.9 (2.0) | 0 |
| **Income decile** | 3.8 (2.1) | 0 |
| **Employment decile** | 4.4 (2.1) | 0 |
| **Education decile** | 5.2 (2.1) | 0 |
| **Eye** |  |  |
| **Right Eye** | 39 [44.3%] | - |
| **Left Eye** | 49 [55.7%] | - |
| **Preop BVA (logMAR)** | 0.3 (0.2) | 0 |
| **Preop Kmax (D)** | 62.5 (9.8) | 8 |
| **Preop K2 (D)** | 53.3 (7.3) | 8 |

*SD*: Standard deviation, *N*: Count (eyes or patients), *IMD*: Index of multiple deprivation, *BVA*: Best visual acuity, *logMAR*: Logarithmic of the minimum angle or resolution, *D*: Dioptre.

**Fig 2.**
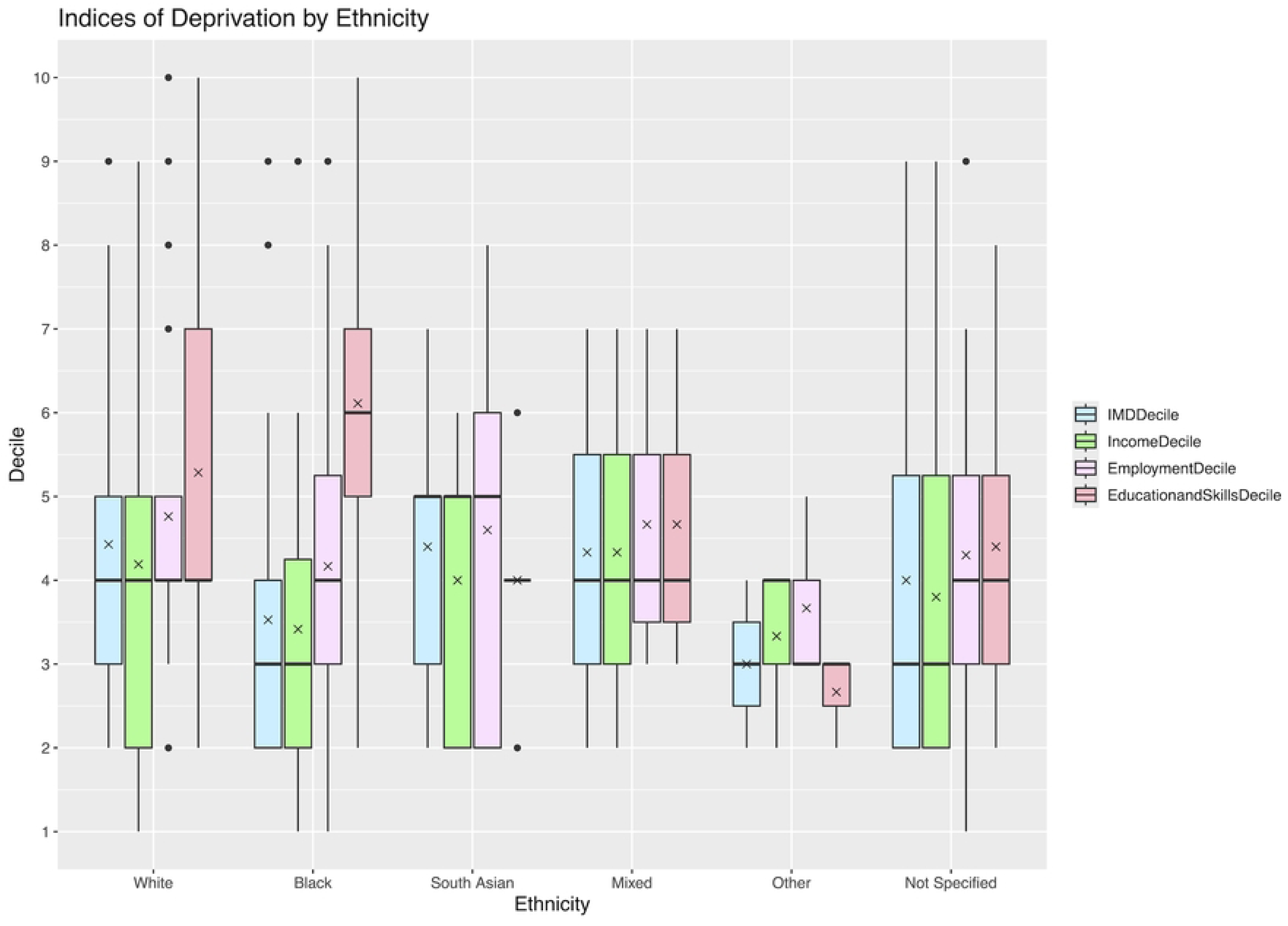
Box plot of ethnicities and deprivation index decile. ‘X’ denotes mean, *IMD*: Index of Multiple Deprivation.

There was an 81.1% (95% confidence interval (CI): 71.2%–87.6%) probability of BVA stability within 52 weeks of CXL (**Fig 3A**). After excluding eight eyes with missing post operative keratometry, there was an 80.7% (67.4%–88.5%) probability of Kmax/K2 stability within 52 weeks of CXL (**Fig 3B**). The probability of non-composite Kmax stability and K2 stability within 52 weeks of CXL was 71.5% (57.0%–81.2%) and 64.6% (49.6%–75.1%) respectively (**Supporting Information**). The time taken for a 50% chance of BVA stability and Kmax/K2 stability was 12.3 weeks and 23.3 weeks respectively. Multivariate Cox-regression analysis showed significant associations (*p*<0.05) between Kmax/K2 stability and the “other” ethnicity group (**Table 2**). No significant associations were identified between VA or corneal topographical stability and other sociodemographic factors. The post-operative complication rate in this cohort was 2.3%.

**Fig 3.**
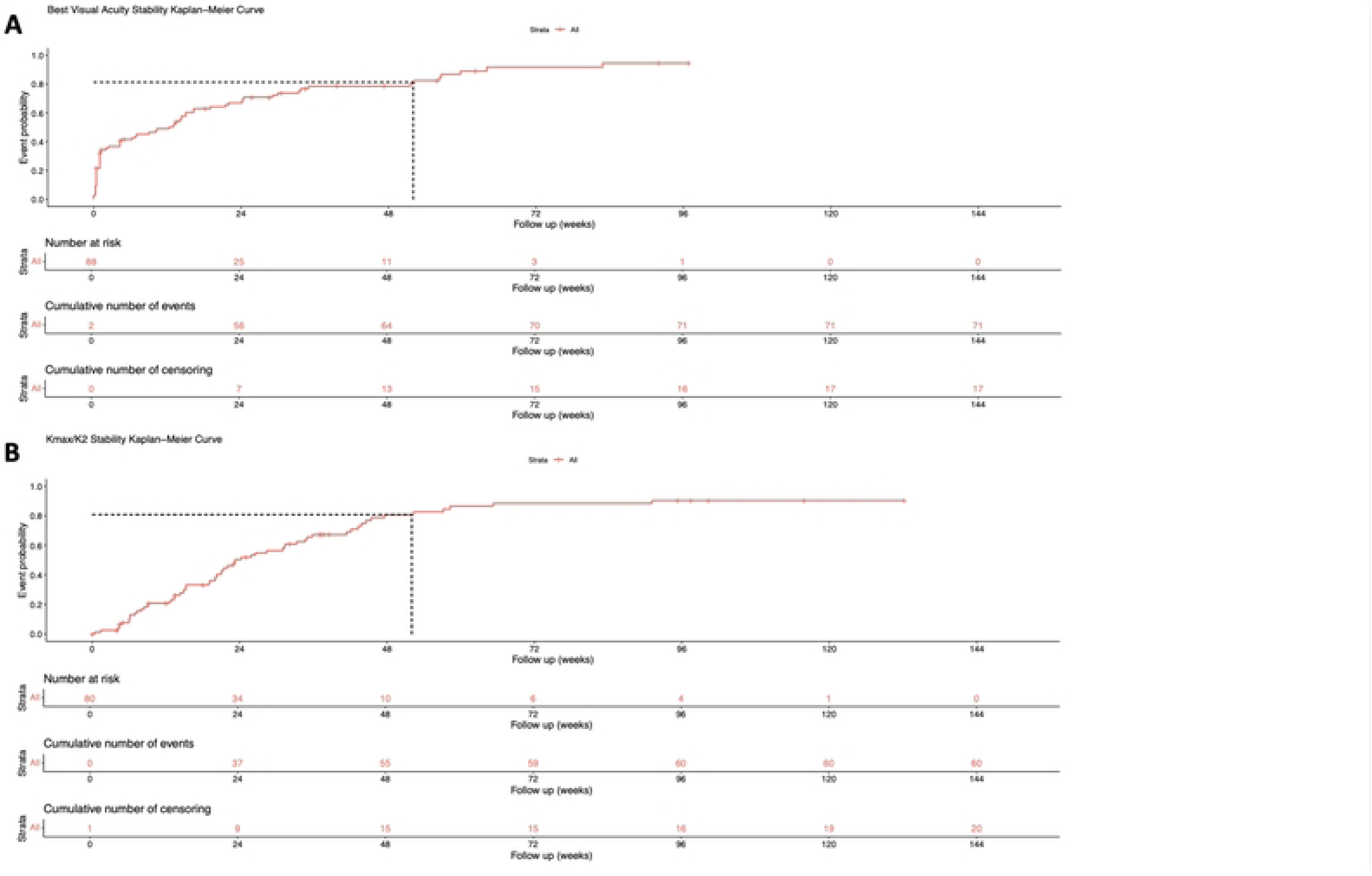
Kaplan-Meier Curves for (A) Best Visual Acuity Stability and (B) Kmax/K2 stability. Dashed line to denote probability of stability at 52 weeks.

**TABLE 2.** MULTIVARIATE COX REGRESSION

|  | Significance | HR | 95% CI for HR |  |
| --- | --- | --- | --- | --- |
|  |  |  | Lower | Upper |
| BVA Stability – Income Decile |  |  |  |  |
| Age | 0.340 | 1.019 | 0.981 | 1.058 |
| Sex - Male | 0.570 | 1.178 | 0.669 | 2.074 |
| Ethnicity - Black | 0.707 | 0.882 | 0.458 | 1.699 |
| Ethnicity - South Asian | 0.651 | 1.305 | 0.412 | 4.140 |
| Ethnicity - Mixed Ethnicity | 0.076 | 3.278 | 0.884 | 12.157 |
| Ethnicity - Other Ethnicity | 0.424 | 1.718 | 0.457 | 6.462 |
| Ethnicity - Not Specified | 0.765 | 0.900 | 0.451 | 1.797 |
| Income Deprivation | 0.877 | 0.991 | 0.883 | 1.112 |
| Preop Kmax (D) | 0.883 | 1.003 | 0.970 | 1.036 |
| Significant Postop Complication | 0.326 | 2.114 | 0.475 | 9.402 |
| BVA Stability – Employment Decile |  |  |  |  |
| Age | 0.342 | 1.019 | 0.981 | 1.058 |
| Sex - Male | 0.609 | 1.159 | 0.659 | 2.038 |
| Ethnicity - Black | 0.657 | 0.861 | 0.445 | 1.666 |
| Ethnicity - South Asian | 0.654 | 1.303 | 0.410 | 4.138 |
| Ethnicity - Mixed Ethnicity | 0.076 | 3.276 | 0.883 | 12.156 |
| Ethnicity - Other Ethnicity | 0.407 | 1.752 | 0.465 | 6.599 |
| Ethnicity - Not Specified | 0.747 | 0.893 | 0.448 | 1.781 |
| Employment Deprivation | 0.609 | 1.033 | 0.919 | 1.155 |
| Preop Kmax (D) | 0.771 | 1.005 | 0.973 | 1.038 |
| Significant Postop Complication | 0.396 | 1.934 | 0.422 | 8.870 |
| BVA Stability – Educational and Skills Decile |  |  |  |  |
| Age | 0.338 | 1.019 | 0.981 | 1.059 |
| Sex - Male | 0.584 | 1.170 | 0.667 | 2.053 |
| Ethnicity - Black | 0.735 | 0.887 | 0.444 | 1.773 |
| Ethnicity - South Asian | 0.663 | 1.295 | 0.405 | 4.142 |
| Ethnicity - Mixed Ethnicity | 0.078 | 3.261 | 0.878 | 12.115 |
| Ethnicity - Other Ethnicity | 0.448 | 1.689 | 0.436 | 6.550 |
| Ethnicity - Not Specified | 0.756 | 0.896 | 0.449 | 1.789 |
| Education and Skills Deprivation | 0.929 | 0.994 | 0.869 | 1.137 |
| Preop Kmax (D) | 0.859 | 1.003 | 0.971 | 1.036 |
| Significant Postop Complication | 0.328 | 2.124 | 0.470 | 9.597 |
| Kmax/K2 Stability – Income Decile |  |  |  |  |
| Age | 0.750 | 1.007 | 0.966 | 1.049 |
| Sex - Male | 0.075 | 1.818 | 0.942 | 3.511 |
| Ethnicity - Black | 0.952 | 1.022 | 0.504 | 2.073 |
| Ethnicity - South Asian | 0.180 | 0.243 | 0.031 | 1.919 |
| Ethnicity - Mixed Ethnicity | 0.982 | 1.018 | 0.220 | 4.701 |
| Ethnicity - Other Ethnicity | 0.011 | 6.558 | 1.545 | 27.834 |
| Ethnicity - Not Specified | 0.671 | 1.186 | 0.540 | 2.608 |
| Income Deprivation | 0.700 | 0.973 | 0.845 | 1.120 |
| Preop Kmax (D) | 0.552 | 1.009 | 0.979 | 1.041 |
| Significant Postop Complication | 0.906 | 1.132 | 0.145 | 8.845 |
| Kmax/K2 Stability – Employment Decile |  |  |  |  |
| Age | 0.693 | 1.009 | 0.967 | 1.052 |
| **Sex - Male** | 0.072 | 1.823 | 0.948 | 3.505 |
| **Ethnicity - Black** | 0.957 | 1.020 | 0.507 | 2.051 |
| **Ethnicity - South Asian** | 0.211 | 0.264 | 0.032 | 2.131 |
| **Ethnicity - Mixed Ethnicity** | 0.969 | 1.031 | 0.224 | 4.749 |
| **Ethnicity - Other Ethnicity** | **0.010** | **6.583** | **0.569** | **27.625** |
| **Ethnicity - Not Specified** | 0.615 | 1.228 | 0.552 | 2.735 |
| **Employment Deprivation** | 0.521 | 0.953 | 0.822 | 1.104 |
| **Preop Kmax (D)** | 0.516 | 1.010 | 0.980 | 1.042 |
| **Significant Postop Complication** | 0.822 | 1.273 | 0.156 | 10.400 |
| **Kmax/K2 Stability – Educational and Skills Decile** |  |  |  |  |
| **Age** | 0.839 | 1.004 | 0.963 | 1.047 |
| **Sex - Male** | 0.650 | 1.847 | 0.962 | 3.545 |
| **Ethnicity - Black** | 0.957 | 1.020 | 0.502 | 2.069 |
| **Ethnicity - South Asian** | 0.162 | 0.231 | 0.030 | 1.805 |
| **Ethnicity - Mixed Ethnicity** | 0.972 | 1.028 | 0.222 | 4.758 |
| **Ethnicity - Other Ethnicity** | **0.008** | **7.658** | **1.717** | **34.163** |
| **Ethnicity - Not Specified** | 0.676 | 1.180 | 0.543 | 2.567 |
| **Education and Skills Deprivation** | 0.603 | 1.038 | 0.903 | 1.193 |
| **Preop Kmax (D)** | 0.553 | 1.009 | 0.979 | 1.041 |
| **Significant Postop Complication** | 0.999 | 0.999 | 0.128 | 7.781 |

*BVA*: Best visual acuity, *CI*: Confidence interval, *HR*: Hazard ratio, *IMD*: Index of multiple deprivation, *D*:
Dioptre.

## DISCUSSION

To the best of our knowledge, this is the first study using survival analysis to investigate the associations between sociodemographic factors, visual acuity and corneal topographic outcomes in keratoconus eyes undergoing CXL. This ethnically and socioeconomically diverse patient cohort offers a unique perspective on the interactions between sociodemographic factors and important clinical outcomes of CXL. In this study, we used multivariate Cox-regression analysis to investigate the associations between sociodemographic factors and CXL outcomes. We identified independent associations between the other ethnicity group and Kmax/K2 stability. However, no associations were identified between other socioeconomic factors, including subdomains of deprivation or other ethnicity groups with the stability of VA or topographical after CXL.

There are limited studies evaluating sociodemographic factors and the progression of keratoconus after CXL. Black and non-white ethnicity, lower income, and unemployment have been associated with a greater severity of keratoconus [26, 28]. Poor parental education in paediatric patients and lower levels of patient education have also been associated with an increased risk of keratoconus progression [33-35], with parental education associated with keratoconus severity [35, 36]. Younger age, allergic conjunctivitis, eye rubbing, high preoperative Kmax have all been associated with keratoconus progression after CXL [37-39].

CXL has been shown to prevent the long-term progression of keratoconus and improve both VA and Kmax in numerous studies [16-22, 40-45]. In a systematic review, Craig et al. reported improvement in VA and Kmax with CXL [46]. At 12 months, the change in logMAR BCVA was -0.09 (95%CI: -0.11; -0.06) across all included publications (n=22) using a random effects model. Meta-analysis of three RCTs (n=3) identified a statistically significant improvement in BCVA of -0.19 (95%CI: -0.26; -0.12) at 12 months with CXL. A -1.03 D (95%CI: -1.34; -0.71) Kmax reduction at 12 months after CXL was reported using a random effects model across all publications (n=18) [46]. RCTs (n=3) that reported on Kmax identified statistically significant reductions of -2.66 D, -1.7 D & -1.45 D at 12 months after CXL [16, 17, 45]. In a study investigating CXL in paediatric patients, an average 3D reduction in K2 was identified at 18 months. Patients have also reported reduced subjective visual symptoms (e.g. glare, halo) and improved visual function (e.g. night driving) after CXL, with subjective symptoms correlating with objective measurements of VA [20].

In our study, we found that that the chance of BVA stability was ∼80% at 12 months. Whilst our survival analyses approach is not directly comparable, it supports prior literature demonstrating the effectiveness of CXL in stabilising VA and preventing the progression of keratoconus. This was further highlighted by the high probability of achieving Kmax/K2 stability at ∼80% at 12 months, which was proportional to the percentage of patients also achieving BVA stability.

The use of Kaplan-Meier survival analysis enables cumulative survival to be estimated using the observational data available from real-world clinical data whilst accounting for censoring. In our study, survival outcomes were measured in patients with two years of follow-up. The study findings therefore provide a useful guidance to keratoconus patients and clinicians on the probability of achieving stable VA over a two-year time period after CXL. The limitations of this study include its retrospective design which can be affected by unadjusted biases, missing data for some patients, and that deprivation is estimated using the IMD which is based on postcode and a small neighbour area (∼1,500 people), which may not reflect individual-level deprivation. Additionally, potential confounding factors associated with keratoconus progression, such as atopy and eye rubbing, were not available. The other ethnicity group had the lowest deprivation status for income, employment and education deprivation domains hence residual confounding may be a cause of the associations found as only one deprivation domain could be adjusted per model due to domain correlations.

In conclusion, survival analysis demonstrated that CXL is associated with the stabilisation of BVA, Kmax and K2 in a significant proportion of eyes over 12 months. Significant associations between other ethnicity group and Kmax/K2 stability were found but no other significant associations were identified between sociodemographic factors, VA and topographic stability after CXL. These findings suggest CXL in patients with keratoconus results in good functional and anatomical outcomes independent of sociodemographic status.

## Data Availability

Approvals to share the dataset is not covered in the audit approval process, the raw data therefore cannot be shared. Programming code is included in the Supporting Information.

## ACKNOWLEDGEMENTS

We would like to acknowledge Magnus Theodorsson, Kimberly Tan and Mohamed Alaaeldin for their contribution to prior cross-linking audits at King’s College Hospital as their data collection methodology helped inspire this study.

## CONTRIBUTORS

Concept: PN, SJ; Data analysis: CH, PN, RG, SJ; Write-up: CH, PN, SJ. Submission: CH, PN, SJ.

## FUNDING STATEMENT

None.

## COMPETING INTERESTS

None of the authors have any conflict of interest to declare.

## PATIENT CONSENT

Not applicable.

## ETHICS APPROVAL

This study was conducted as an audit of the local outcomes after CXL for KC using the published literature [22] as the gold standard. The study used anonymised retrospective data and, as an audit, was exempt from research ethics committee approval. The audit was registered and approved by the Ophthalmology Care Group Audit Lead at King’s College Hospital on the 23rd October 2023 and the audit ID was AS008.

## SUPPORTING INFORMATION

**S1 Fig. Best visual acuity stability after corneal cross-linking.**

**S2 Fig. Kmax/K2 stability after corneal cross-linking.**

**S3 Fig. Kmax stability after corneal cross-linking.**

**S4 Fig. K2 stability after corneal cross-linking.**

## Notes

### Competing Interest Statement

The authors have declared no competing interest.

### Funding Statement

The author(s) received no specific funding for this work.

